# Fox Insight Collects Online, Longitudinal Patient-Reported Outcomes and Genetic Data on Parkinson’s disease

**DOI:** 10.1101/19002659

**Authors:** Luba Smolensky, Ninad Amondikar, Karen Crawford, Scott Neu, Catherine M. Kopil, Margaret Daeschler, Lindsey Riley, 23andMe Research Team, Ethan Brown, Arthur W. Toga, Caroline Tanner

## Abstract

Fox Insight is an online, longitudinal health study of people with and without Parkinson’s disease with targeted enrollment set to at least 125,000 individuals. Fox Insight data is a rich data set facilitating discovery, validation, and reproducibility in Parkinson’s disease research. The dataset is generated through routine longitudinal assessments (health and medical questionnaires evaluated at regular cycles), one-time questionnaires about environmental exposure and healthcare preferences, and genetic data collection.

Qualified Researchers can explore, analyze, and download patient-reported outcomes (PROs) data and Parkinson’s disease-related genetic variants at https://foxden.michaeljfox.org. The full Fox Insight genetic data set, including approximately 600,000 single nucleotide polymorphisms (SNPs), can be requested separately with institutional review and are described outside of this data descriptor.

Fox Insight is sponsored by The Michael J. Fox Foundation for Parkinson’s Research.

## Background & Summary

Parkinson’s disease (PD) is the second most common neurodegenerative disease, with prevalence expected to increase over time [1, 2]. Parkinson’s disease presents with a wide range of manifestations; motor symptoms, non-motor symptoms, response to medication, and variable rate of progression among those affected. This variability has introduced challenges in understanding disease progression, clarifying underlying pathophysiology, providing meaningful treatments, and fully grasping which symptoms are most detrimental to patients. In-person trials classically enroll participants who already have access to specialist care, with milder symptomatology, better cognition, and less diversity than the general population [3, 4]. As a result, observational studies with larger sample sizes, longer follow-up, and deeper patient perspective are needed to improve our disease understanding.

Online data collection offers a mechanism to address these research challenges. Online surveys may pose less subject burden, and enrollment can target traditionally underrepresented demographics. Virtual data collection is effectively employed in other settings to achieve large sample sizes and facilitate data access and analysis, such as in the National Institute of Health’s *All of Us* Research Program [5]. Internet usage among those over 65, the population most likely to develop Parkinson’s disease, has risen substantially in the last several years, with 67% reporting regular internet usage [6]. With the blend of growing internet usage, participation in online research studies, and digitized self-reported assessments, Parkinson’s disease lends itself well to online data collection.

In addition, genetic variation is thought to play a significant role in Parkinson’s disease etiology likely in concert with environmental exposure [7]. In a minority of cases, a rare single gene mutation is strongly associated with Parkinson’s disease. Other mutations increase risk but have lower penetrance [8]. Multiple genetic variants have been aggregated into a genetic risk score and combined with phenotypic characteristics to classify people with our without Parkinson’s disease [9]. Remotely assessed self-reported genotype and phenotype information suggested different clinical subtypes in one online study [10]. Genetic variation and risk alleles are an important component to understanding many aspects of Parkinson’s disease, and genetic data is a large asset.

Fox Insight is an online study consisting of regularly-administered questionnaires collected longitudinally over several years. Fox Insight is open to participants with self-reported PD and those without self-reported PD (caregivers and healthy controls). Fox Insight data is immensely useful in improving understanding of patient experiences and complementing PROs with Parkinson’s disease genetic risks and modifiers.

Fox Insight integrates validated PRO instruments and PD-related questionnaires as illustrated in Figure 1. The content and cadence of each questionnaire is dependent on participant self-reported diagnosis. Fox Insight also includes the implementation of one-time questionnaires and genetic data collection. By design, Fox Insight can support modifications to multi-modal data collection in alignment with evolutions in Parkinson’s disease research. This flexibility is enabled by Fox Insight’s infrastructure, an agile-developed web application, built through a software development framework that emphasizes phased deployment, that manages enrollment, e-consent, and a collection of routine longitudinal assessments.

**Figure 1:**
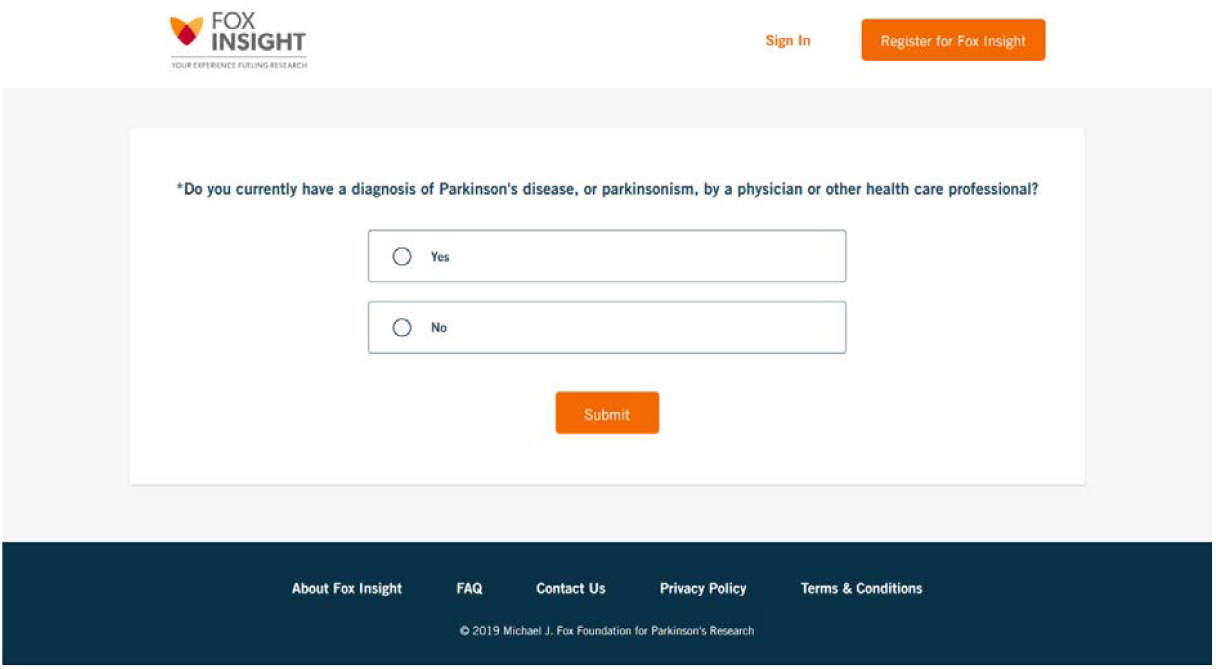
Online questionnaires. Figure 1 demonstrates the style in which online questionnaires are presented to participants in Fox Insight. This Fox Insight screenshot shows how participants select self-reported Parkinson’s disease diagnosis.

## Methods

Fox Insight is open to participants, aged 18 or older, who provide informed consent through the Fox Insight website; informed content and study protocol are reviewed by the New England IRB (IRB#: 120160179, Legacy IRB#: 14-236, Sponsor Protocol Number: 1, Study Title: Fox Insight). Volunteers are recruited through digital channels (e.g. social network ads, search engine marketing, and email newsletters) and on-the-ground recruitment efforts (e.g. research events, clinician referrals). Upon registration, participants are divided into two primary cohorts, those with Parkinson’s disease and those without. Importantly, participants without PD are asked about new diagnoses every three months, and are given a different set of assessments based on self-reported Parkinson’s disease diagnosis. People with Parkinson’s disease respond to health, non-motor assessments, motor assessments, quality of life, and lifestyle questionnaires (through twenty questionnaires that are part of each routine longitudinal assessment). In contrast, people without Parkinson’s disease respond only to health and lifestyle questionnaires (through a separate grouping of thirteen questionnaires in each routine longitudinal assessment). Participants that meet the pre-set eligibility criteria of optional, one-time questionnaires are invited to participate in additional PRO collection. People with Parkinson’s disease based in the US who have completed at least twenty questionnaires in a routine longitudinal assessment are invited to participate in genetic research.

The following methods describes the three data acquisition sources of Fox Insight: routine longitudinal assessments, one-time questionnaires, and genetics as illustrated in Figure 2. Routine longitudinal assessments form the main study activities and are collected through a custom survey application developed by Mondo Robot [12], a creative digital agency. One-time questionnaires are deployed through Qualtrics® survey software [13], leveraged for additional survey programming rules. Finally, genetic data are collected in collaboration with 23andMe, Inc. [14], a personal genetics company.

**Figure 2:**
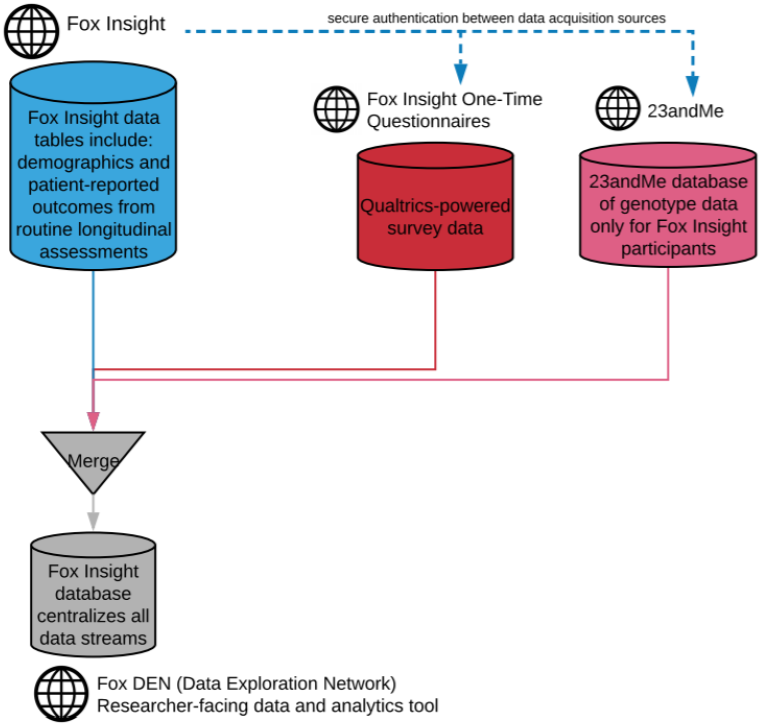
Fox Insight Data flow. Figure 2 represents the data flow in Fox Insight combining patient-reported outcomes and genetic data into Fox Insight’s data ecosystem. Demographic data and patient-reported outcomes from routine longitudinal assessments are merged with responses from one-time questionnaires and genetic data into a central database accessible to researchers.

### Routine Longitudinal Assessments

Routine longitudinal assessments are hosted through an online survey platform and offered to participants based on self-reported Parkinson’s disease diagnosis. The assessment schedule is derived from the participant’s registration date. These assessments aim to comprehensively evaluate many potential aspects in Parkinson’s disease, including motor impairment, non-motor symptoms, medication efficacy, functional impact, and quality of life. Validated instruments are used, when possible, such as the Movement Disorders Society – Unified Parkinson’s disease Rating Scale (MDS-UPDRS) Part II, the Non-Motor Symptoms Questionnaire (NMSQUEST), and the Geriatric Depression Scale (GDS), among others (Table 1).

**Table 1:**
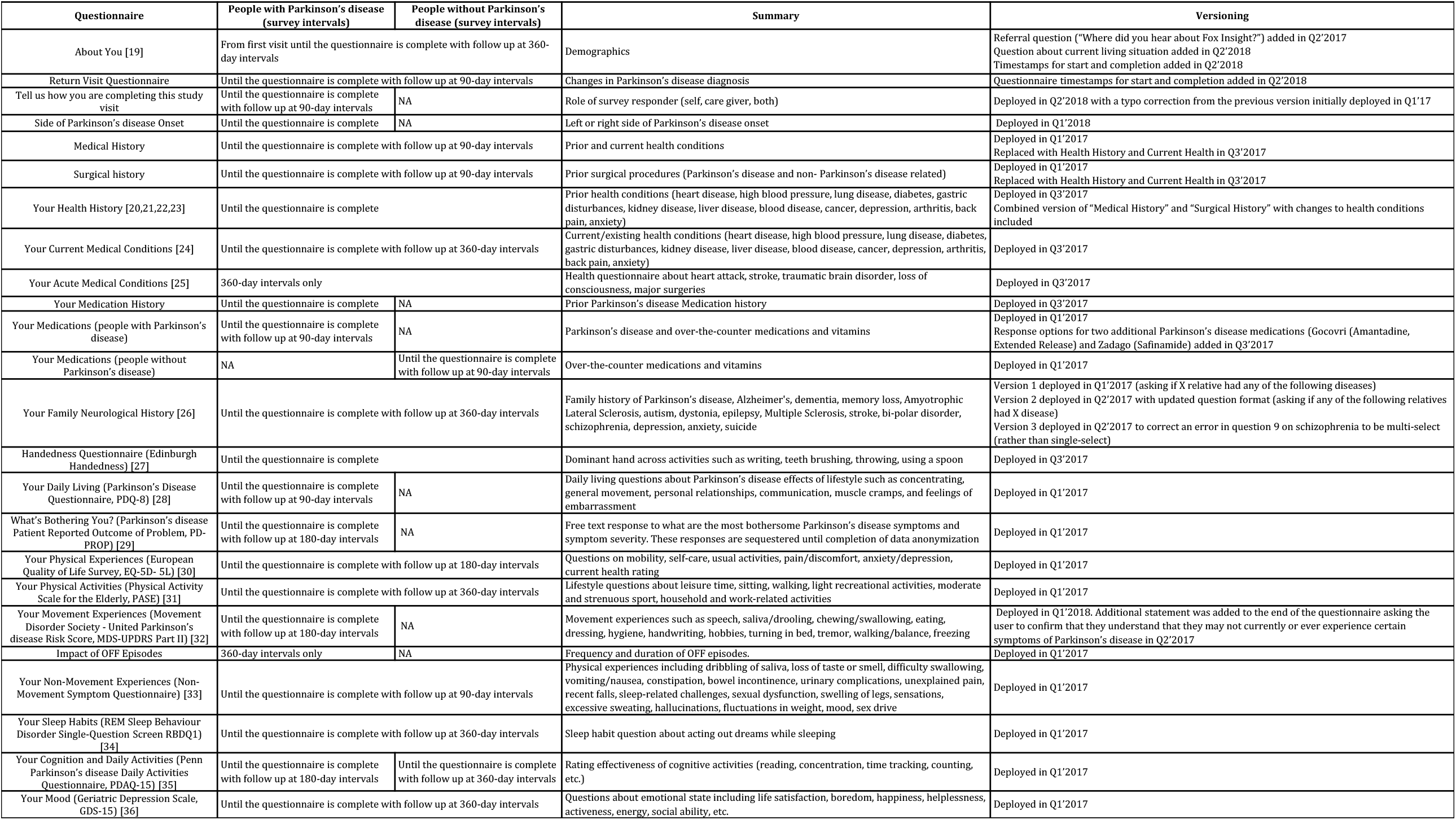
Routine Longitudinal Assessments. Table 1 summarizes questionnaires offered to Fox Insight participants, the cohort, the associated frequency (which may be subject to change as the study evolves), and questionnaire versioning. Questionnaires integrate a battery of assessments that comprehensively evaluates motor and non-motor symptoms using standardized instruments when available, with the official names listed in parentheses.

Data collection from routine longitudinal assessments is governed by survey logic. More specifically, this includes:

1. Participants who answer “Yes” to the registration question “Do you currently have a diagnosis of Parkinson’s disease, or Parkinsonism, by a physician or other health care professional?” are presented with Parkinson’s disease assessments. Those who answer “No” are classified as people without Parkinson’s disease and receive a different set of questionnaires.
2. Questionnaires are presented sequentially; a participant cannot begin a second questionnaire without completing the first.
3. Participants cannot explicitly skip questions within an opened questionnaire and can instead respond “Prefer Not to Answer” to move onto the next question. The only empty values collected in routine longitudinal assessment data are from bifurcated logic, incomplete surveys, or undistributed questions.
4. Sets of questionnaires are repeated at regularly recurring intervals (Table 1) at which time, a participant is invited, via email, to answer these assessments in Fox Insight.
5. Participants can update Parkinson’s disease diagnosis, living situation, and hospital experience every three months in Fox Insight. If a participant indicates a change in diagnosis, the participant is redirected to a new, alternate set of questionnaires consistent with the change in Parkinson’s disease diagnosis to best capture current health, including a full baseline battery for newly diagnosed Parkinson’s disease. Subsequent routine longitudinal assessments continue to be based on the updated diagnosis and initial study registration date.
6. Assessments can be modified, added, or removed. A participant sees changes to available questionnaires at the start of the next complete assessment interval.
7. Responses to a survey question can determine the deployment and collection of another related survey question. Condition-based questions that are not presented to participants have empty values in the data set. For instance, if a participant answers “Have you ever had a form of heart disease?” in the affirmative, then the following question asks “What kind of heart disease did you have?” and the participant selects from a drop down list of heart disease options. An initial answer of “No, I have not had a form of heart disease” skips the second, follow up question and the response values are empty in the output dataset.
8. Participants can review a summary of responses to an individual questionnaire and can change a question response ahead of finalizing questionnaire submission. In addition, questionnaire responses can be reviewed/revised at any point before the participant receives the next set of assessments.

### One-Time Questionnaires

One-time questionnaires (Table 2) are deployed through Fox Insight to enrich the PRO data collected through routine longitudinal assessments with additional validated instruments. These questionnaires can collect cross-sectional data from novel or unique instruments not included in routine longitudinal assessments. For instance, one-time questionnaires can be a useful first step for in-person trials as a means of obtaining patient perspective during research development, evaluating interest in specific interventions, or targeting recruitment in clinical trials. The ability to deploy one-time questionnaires is an enormous advantage of the Fox Insight platform. The frequency and content of questionnaires is vetted by study leadership to ensure alignment with scientific goals.

**Table 2:** One-Time Questionnaires. Table 2 summarizes scope and eligibility criteria of one-time questionnaires offered in Fox Insight (which may be subject to change as the study evolves).

| Survey | Summary | Eligibility Criteria |
| --- | --- | --- |
| Environmental Exposure Questionnaires (PD-RFQ-U)<br>• Alcohol<br>• Caffeine<br>• Smoking and Tobacco<br>• Head Injury and Concussion<br>• Pesticides at Work<br>• Pesticides in Non-Work Settings<br>• Residential History<br>• Physical Activity and Sleep<br>• Height and Weight<br>• Calcium Channel Blocker Medication History<br>• Anti-Inflammatory Medication History<br>• Occupation<br>• Toxicant<br>• Female Health History | • Detailed questions about lifestyle, personal habits, living and work environments, medication and healthy history<br>• This survey is continuously recruiting. | All participants |
| Impact and Communication About OFF Periods | • Asks care partners and patients to describe how they discuss OFF<br>• This survey is designed by Connie Marras and active recruitment spanned from 02-07-2018 to 03-30-2018. |  |
| The Financial and Social Impact of Parkinson's disease Survey | • Financial, accounting, and tax-related questions on health and medical spending related to Parkinson's disease to understand the economic burden of the disease<br>• This survey is designed by the Lewin Group and beta tester recruitment spanned from 09-16-2018 to 10-16-2018. | All US-based participants |
| Patient Therapeutic Preferences Questionnaire by the Medical Device Innovation Consortium (MDIC) | • Responses to hypothetical medical situations and procedures to determine patient preferences to inform FDA processes for medical device review<br>• This survey is created by MDIC and active recruitment spanned from 11-27-2017 to 01-12-2018. | People with Parkinson's disease |
| Understanding OFF and ON in Parkinson's disease Patients | • Explores how patients experience, and communicate about OFF and ON periods associated with Parkinson's disease<br>• This survey is created by the Parkinson's disease Education Consortium and active from 11-26-2018 to 12-11-2018. | People with Parkinson's disease in the US who report taking at least one Parkinson's disease-related medication |

<sup>1</sup> These authors contributed equally to this work.
<sup>2</sup> These authors jointly supervised this work.

### Fox Insight Genetic Data

Genotyping, through 23andMe, will be available for up to 17,000 participants with Parkinson’s disease in the US who have completed a series of routine longitudinal assessments (5,000 participants have been genotyped at the time of this Data Descriptor). This eligibility criteria of requiring phenotypic data collection upfront ensures valuable context for interpreting and analyzing genotype data; more so, researchers can explore correlations between genetic variations and phenotypic manifestations. Eligible participants provide a sample using 23andMe’s saliva collection kit. Samples have been genotyped on a variety of genotyping platforms. Within Fox Insight, 6.9% of participants are genotyped on the V3 platform which is based on the Illumina OmniExpress + BeadChip and contains a total of about 950,000 SNPs, 12.7% of participants are genotyped on the V4 platform which is a fully custom array of about 570,000 SNPs, and 80.4% of participants are genotyped on the V5 platform which is in current use and is a customized Illumina Infinium Global Screening Array of about 690,000 SNPs. As part of the resulting dataset, several genetic variants that may be relevant for Parkinson’s disease research and have a non-identifiable prevalence within the Fox Insight cohort (including variants located near GBA, LRRK2, APOE, PRKN, MCCC1, BIN3, and the HLA locus) are available in tabular form alongside phenotypic data in Fox Insight’s public repository. These variants are included as categorical data to democratize data access and interpretation for otherwise complex SNP output (the full set of SNPs is available upon request to qualified researchers).

### Data Centralization

Participant answers to routine longitudinal assessments, one-time questionnaires and genetic data from key variants are integrated in a public repository managed at the USC Laboratory of Neuro Imaging, Mark and Mary Stevens Neuroimaging and Informatics Institute. Using dates of birth provided during user registration, dates associated with participant answers are converted to participant ages to protect patient confidentiality. As questions for a single routine longitudinal assessment may be edited and answered intermittently, the total number of days used to complete each survey is also recorded for each participant. Along with dates of birth, unrestricted and free form textual answers are quarantined from the general public data set; when appropriate, “derived” variables are defined for those questions to filter out (e.g., reject non-decimal number values) arbitrary (and possibly patient-identifying) responses. Derived variables are also added for cases in which participants are allowed to answer a question in different ways (e.g., enter weight in pounds or kilograms) in order to help standardize these responses.

## Code availability

Fox Insight is built by several technology partners, each with its own policies on code availability. Routine longitudinal assessments are developed through a web-based application built on Ruby on Rails® [15] software by Mondo Robot and the code base is proprietary. One-time questionnaires are deployed through Qualtrics®; while the survey platform code is proprietary, Qualtrics® provides an open source application programming interface (API) for data processing [16]. SQL code, developed at the Laboratory of Neuro Imaging, used to collate and process data is proprietary.

## Data Records

Data collected from each survey is aggregated into a single table and is available via a comma separated value (CSV) file. Variable values are encoded according to a data dictionary, which accompanies each download from Fox Insight Data Exploration Network (Fox DEN) at https://foxden.michaeljfox.org (access and usage notes detailed in later sections). Participant ages are provided alongside time-dependent data. Additional metrics (e.g., variable vectors per subject recording data availability, histograms of variable values) are pre-computed to facilitate searching and data grouping by researchers. Data from multiple surveys may be dynamically combined into a single table for downloading using Fox DEN. A pre-selected set of 18 SNPs is available in tabular format complemented by genetic metadata including genotype no-call rate and genotype chip version. The data dictionary (Table 4) describes metadata for the collected variables for each survey question in the routine longitudinal assessments and one-time questionnaires. The complete data dictionary, of 2,000 collected variables, is available for download in Fox DEN.

**Table 4:**
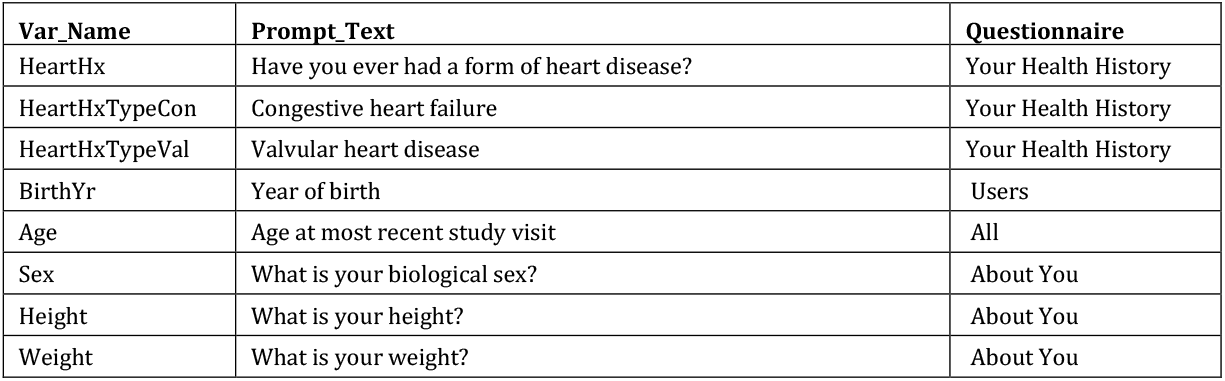
Data Dictionary for Fox Insight Assessments. Table 4 provides a snippet of the full data dictionary demonstrating variable truncation, corresponding questionnaire, and code names.

## Technical Validation

Technical Validation for Fox Insight is bifurcated into tool and data validation. Data validation closely reviews caveats associated with collecting patient reported outcomes and compares sex chromosome to self-reported sex for genetic data validation.

### Deployment of Routine Longitudinal Assessments

To verify the appropriate deployment of routine longitudinal assessments, development tests are routinely conducted by Mondo Robot. Using RSpec [17], a testing framework for Ruby on Rails®, unit tests are run on isolated pieces of code functionality. These unit test include, but are not limited to, database querying for cadence expiration and questionnaire assignment based on registration date. All unit tests automatically run when code is moved into development, staging, and production environments.

While platform tests verify that questionnaires are deployed according to set intervals, post-tests spot check data collection nuances from said tools. For example, data from the Physical Activity Scale for the Elderly (PASE) assessment is expected to be collected regularly. There are 21,484 participants (as of 01-24-2019) who completed the questionnaire in the first round of longitudinal assessments and 285 (1.32% of total) who skipped this assessment entirely in the first set of routine longitudinal assessments. Fox Insight successfully deploys the PASE questionnaire to participants who skip the questionnaire in subsequent assessment periods until a complete questionnaire is submitted; in fact, three-quarters (127) of the participants who skipped PASE in the initial battery of assessments go on to complete the survey in the subsequent assessment period. Redeploying incomplete assessments helps establish a more robust PRO data set.

### Collected Data

The aforementioned data collection methods converge to form a large sample size of PROs from routine longitudinal assessments, one-time questionnaires, and genetic data as illustrated in Table 3.

**Table 3.** Demographics and Collected Data in Fox Insight. Table 3 highlights the scale of collected data in Fox Insight and key cohort characteristics. As of Q1’19, there are over 22,000 people with Parkinson’s disease enrolled making Fox Insight the largest prospectively followed Parkinson’s disease cohort worldwide, exceeding the second largest cohort of 12K people with Parkinson’s disease followed in the Parkinson’s disease Outcome Project [18]. Of the 30,436 total individuals enrolled in Fox Insight, 72.9% (n = 22,205) participants are people with Parkinson’s disease. The average age of the Parkinson’s disease cohort is 66 and these participants, on average age, have been diagnosed for over 6 years. At the time of this Data Descriptor, the Fox Insight dataset has a larger sample size of cross-sectional data than longitudinal data; 90.5% (n = 20,099) of people with Parkinson’s disease have answered at least one questionnaire and 47.7% (n = 10,600) of people with Parkinson’s disease participants have continued participating in routine longitudinal assessments. People without Parkinson’s disease exhibit a similar trend in assessment completion. Optional one-time questionnaires are completed by a comparatively lower proportion of the study population with 34.8% (n = 7,726) of people with Parkinson’s disease, and 14.2% (n = 1,174) of people without Parkinson’s disease participating in one-time surveys. As of 03-06-2019, 5,880 total participants agreed to genetic data collection and 5,092 participants are genotyped.

| Full Cohort (N = 30,436) | People with Parkinson's disease | People without Parkinson's disease |
| --- | --- | --- |
| Total enrolled N (% of total) | 22,205 (72.9%) | 8,231 (27.1%) |
| Age* (mean (sd) years) | 65.87 (10.08) | 56.67 (14.07) |
| Sex |  |  |
| Female | 8,683 (38.9%) | 5,501 (66.8%) |
| Male | 10,813 (48.7%) | 1,705 (20.7%) |
| Length of Parkinson's disease diagnosis* (mean (sd) years) | 6.61 (5.88) | NA |
| Full Data Collection |  |  |
| First completion of routine longitudinal assessments: Cross-sectional |  |  |
| Participants (number, percent of total) | 20,099 (90.5%) | 7,198 (87.4%) |
| Questionnaires (volume, average per participant in first assessment) | 252,079 (11.35) | 65,027 (7.9) |
| Routine longitudinal assessments: Longitudinal |  |  |
| Participants (number, percent of total) | 10,600 (47.7%) | 2,664 (32.3%) |
| Questionnaires (volume, average per participant in a subsequent assessments) | 239,461 (10.8) | 42,951 (5.2) |
| Repeat assessments per participant (mean (sd)) | 3.7 (2.1) | 3.1 (2.1) |
| Ancillary Surveys |  |  |
| Participants (number, percent of total) | 7,726 (34.8%) | 1,174 (14.4%) |
| Questionnaires (volume, average per participant) | 50,449 (2.2) | 6,527 (0.8) |
| Genetic Sub-Study |  |  |
| Total enrolled (genotype completed) | 5,880 (5,092) | NA |

*Note: Cross sectional refers to the first battery of assessments as part of routine longitudinal assessments. 'Age' and 'Length of Parkinson's disease Diagnosis' are calculated from time of Fox Insight registration. PRO data as of 01-24-2019. Enrollment in the Genetic Sub-Study as of 03-06-2019.*

### Beta Participants

Approximately 16% of total participants (N= 4,697) are part of Fox Insight’s beta group, defined as those joining before the March 2017 soft launch of Fox Insight. Responses to routine longitudinal assessments for all beta group participants are included in the Fox Insight data set. Data from the beta group could be subject to questionnaire versioning and inconsistencies associated with platform troubleshooting and optimization.

### Missing Data

As a comment on missing data collection, there are 2,868 (as of 01-24-2019) participants who did not complete demographic questions in About You; a subset of ∼500 individuals skipped this questionnaire due to a platform glitch which has been resolved as of Q3’2017. Participant drop-off also results in missing demographic data.

There are 1,476 participants (as of 01-24-2019) who have two consecutive assessment periods starting on the same day (i.e., questionnaire responses are associated with the same “Days since Acquired” variable). This questionnaire assignment error has since been fixed. The resulting output for these participants includes data from the most recent, later, routine longitudinal assessment; data from former assessments are skipped.

As routine longitudinal assessments are completed sequentially, there is observed drop-off from the first to the last assessment within the same period of approximately 10.1%.

### Validating Fox Insight Genetic Data

The sex chromosome and self-reported sex match for 99.76% of the genetic sub-study participants. As additional validation documentation, tables of genotyping call rates are provided. The genotyping rates are ancestry and genotyping platform specific and are derived from the 23andMe participant database (i.e. the table for genotyping rates of participants with European ancestry genotyped on the V5 platform was computed on 23andMe participants with European ancestry genotyped on the V5 platform).

## Usage Notes

### Fox DEN User Interface

Using the Fox DEN interface, investigators may explore, select data, and apply statistical methods using user-created cohorts based on subject demographics, PROs, and SNPs. Routine longitudinal assessments, one-time questionnaires, and genetic data are organized in a tree structure. The tree is filtered using drop-down categories (e.g., questionnaires, genetic data) or keyword searches. The distributions of participants’ questionnaire responses and SNP variants are visualized when selected in the tree. Categorical variables can be reduced to user-defined binary variables, which are useful inputs to the statistical methods. Variable visualizations are dependent upon the user-selected cohort, and this provides visualizations specific to subsets of participants. Cohorts are created by recursively selecting values of a variable and using them as a filter to subset a parent cohort. Cohorts are viewed in a tree structure that shows how the cohorts are inherited from one another as well as the filters that define them. Fox DEN supports common statistical methods (linear correlation, logistic regression, chi-square and T-test) through drag and drop operations of its cohorts and variables. A “Guided Statistics” wizard provides step-by-step guidance in choosing appropriate statistical methods for user selections.

### Access

To access Fox Insight data through the Fox DEN tool, researchers are asked to complete and e-sign a data use agreement at https://foxden.michaeljfox.org. There are two sets of data use agreements; the first allows researchers to access responses from routine longitudinal assessments, one-time questionnaires, and pre-selected Parkinson’s disease-related genetic variants. Separately, the second data use agreement allows researchers, with institutional review, to request access to all SNPs. Data dictionaries and genetic data documentation are available in Fox DEN as reference guides.

Researchers can register for an account through Fox DEN and upon successful completion of the Fox Insight data use agreement, researchers can explore, analyze, and download data as illustrated in Figure 3.

**Figure 3:**
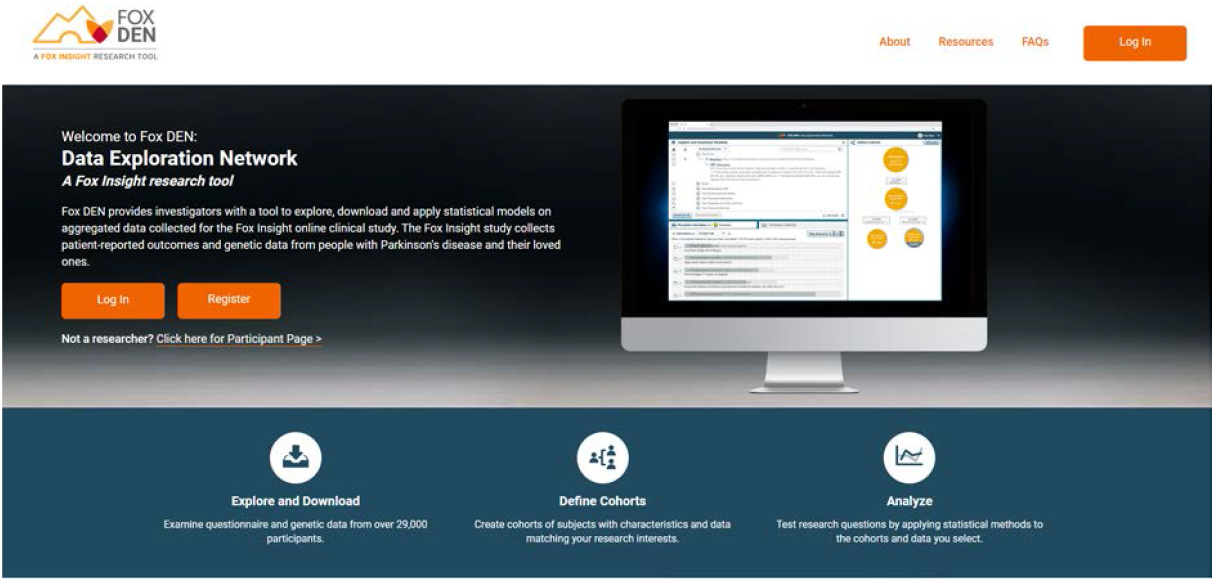
Fox DEN. Figure 3 shows a screenshot of the Fox Den home page. Qualified researchers can register for Fox Insight data access through the home page or log in with approved credentials to explore and download data.

## Data Availability

Fox Insight data can be accessed through the Fox DEN tool. Researchers can complete and e-sign a data use agreement at https://foxden.michaeljfox.org.

## Acknowledgements

The authors thank the 30K+ Fox Insight participants who volunteered their time, responses, and dedication to the study. We also thank The Michael J. Fox Foundation for Parkinson’s Research for sponsoring Fox Insight as well as the Foundation staff who have supported the study, particularly Sohini Chowdhury, Deputy CEO, and Todd Sherer, CEO, who have championed Fox Insight from its inception. In addition, the authors would like to thank the Fox Insight team, Marketing, Communications, Development and Research departments for all their diligent support including: Lauren Bataille, Allison Boiles, Debi Brooks, Bradford Casey, Kristin Demafeliz, Liz Diemer, Rachel Dolhun, Veronique Enos Kaefer, Allyse Falce, Brian Fiske, Mark Frasier, Michele Golombuski, Stephen Gradinscak, Jamie Hamilton, Samantha Hutton, Andrea Katz, Sean Keating, Krishna Knabe, Ashwin Mallya, Lauren McLaughlin, Amanda Melnick, Sherri Mosovsky, Emily Moyer, Gwen Schroder, Bernadette Siddiqi, Ariella Silberstien, Alisha Steindecker, Stephen Streicher, Shruti Suresh, Kristen Teesdale, Holly Teichholtz.

The authors thank Fox Insight leadership team and Fox Insight Executive Steering Committee for ongoing scientific counsel, including: Lana Chahine, Marissa Dean, Roseanne Dobkin, Ken Marek, Connie Marras, Ira Shaulson, David Standaert and Monica Korell as the global project manager of Fox Insight.

We also acknowledge 23andMe for partnering with Fox Insight on genetic data collection, quality control, and data sharing and would like to thank the research participants and employees of 23andMe for making this work possible. Members of the 23andMe Research Team are: Michelle Agee, Babak Alipanahi, Adam Auton, Robert K. Bell, Katarzyna Bryc, Paul Cannon, Sarah Clarke, Sarah L. Elson, Peter Fonseca, Pierre Fontanillas, Nicholas A. Furlotte, Barry Hicks, David A. Hinds, Karl Heilbron, Karen E. Huber, Ethan M. Jewett, Yunxuan Jiang, Aaron Kleinman, Keng-Han Lin, Nadia K. Litterman, Marie Luff, Matthew H. McIntyre, Kimberly F. McManus, Joanna L. Mountain, Elizabeth S. Noblin, Carrie A.M. Northover, Steven J. Pitts, G. David Poznik, Helen M. Rowbotham, J. Fah Sathirapongsasuti, Madeleine Schloetter, Janie F. Shelton, Suyash Shringarpure, Chao Tian, Joyce Y. Tung, Vladimir Vacic, Xin Wang, Catherine H. Wilson, Anne Wojcicki, Linda P.C. Yu.

The authors are deeply grateful to the technology partners who have been instrumental in building and scaling Fox Insight. The authors also thank Mondo Robot for developing and maintaining the Fox Insight platform specifically Jesse Manning, the lead developer at Mondo Robot who has been instrumental in building data capture mechanisms and Shawn Cimock, Matt Fender, Ben Frederick, Kristian Hansen, Chris Hess, Jon McKinney, Kai Raider, and Britt Winn at Mondo for managing Fox Insight projects. We’d like to thank the Laboratory of Neuro Imaging at the University of Southern California for centralizing and managing Fox Insight and developing Fox DEN. Special thank you to Viktoria Andreeva, Laura Brovold, and Leonya Ivanov from Rancho Biosciences for quality control and technical consulting on genetic data.

We’d like to extend a warm thank you to Fox Insight’s statistical partners at the University of Iowa, specifically Janel Barnes, Chelsea Caspell-Garcia, Chris Coffey, Dixie Ecklund, Traci Schwieger, and Maggie Spencer for creating the data dictionary and analysing Fox Insight data as well as Blackfynn, specifically Chris Baglieri, Iris Chin, Amanda Christini, Leo Guerico, Mark Hollenbeck, Eva von Weltin for building analytics dashboards and providing statistical consultation to the Fox Insight Executive Steering Committee.

The Fox Insight team thanks and acknowledges Qualtrics for developing the one-time questionnaire survey application.

## Author contributions

L.S. conceptualized, wrote, synthesized data, and managed the collaborative development of the manuscript. N.A. analysed data and technically validated data output. A.R., K.C. and S.N. wrote data centralization components, oversaw data management, and developed Fox DEN. K.K. conceptualized data collection and study design as well as heavily reviewed manuscript drafts. M.D. implemented and maintained questionnaire deployment and study design operations. L.R. project managed clinical operations for the study. The 23andMe Research Team collected the genetic data, and wrote and edited genetic data components of the manuscript. E.B. wrote and edited clinical background, study relevance, and study design. C.T. revised manuscript.

